# New framework for the surveillance and early warning of influenza: fixed individuals regular reporting mechanism

**DOI:** 10.1101/2024.10.27.24316208

**Authors:** Xiaoyue Sun, Hongwei Wang, Gongke Yang, Ruifang Guo, Shan Zhang, Ruiyu Chai, Shifang Qu, Siyu Liu

**Affiliations:** Department of Epidemiology and Biostatistics, School of Public Health, Jilin University, Changchun 130021, China; State Key Laboratory for Diagnosis and Treatment of Severe Zoonotic Infectious Diseases, Key Laboratory for Zoonosis Research of the Ministry of Education, Jilin University, Changchun 130021, China

**Author notes:** Correspondence to: Siyu Liu, PhD. These authors contributed equally to this work (Xiaoyue Sun, Hongwei Wang). **Author Contributions** S.L.: Conception and design, Method, Revised the language/article. X.S.: Collection and assembly of data, Data analysis and interpretation, Manuscript writing. H.W.: Collection and assembly of data, Data analysis and interpretation, Manuscript writing. G.Y.: Manuscript writing, Collection and assembly of data. S.Z.: Revised the language/article. R.G.: Collection and assembly of data, Revised the language/article. R.C.: Collection and assembly of data. S.Q: Collection and assembly of data. **Competing Interest Statement** The authors declare that they have no competing interests.

**Keywords:** influenza surveillance, fixed individuals, infectious disease, cohort study

## Abstract

Influenza is an acute respiratory infection caused by the influenza virus. Influenza is not only a major burden on human health, but also a major public health challenge, so it is very necessary to conduct surveillance and early warning of influenza. However, the existing monitoring system is mainly based on sentinel monitoring, which has some limitations in information feedback and reliability. Other new monitoring systems also have shortcomings such as insufficient representation and comprehensive coverage. Therefore, we propose a regional influenza surveillance method based on fixed individuals. This method refers to the epidemic characteristics of influenza, selects representative fixed monitoring individuals, and makes them directly upload their physical conditions on a regular basis to judge the occurrence or not of influenza, and determines the judgment method of the severity of influenza. Our proposed method can detect influenza timely and accurately and give early warning, and make more effective use of health resources, which is of great significance for the development of influenza surveillance system. In addition, the monitoring of influenza will play an important role in the monitoring and early warning of new infectious diseases. Importantly, the surveillance method based on fixed individuals can provide a theoretical basis for the cross-sectional study of infectious diseases and make up the gap in the cohort study of infectious diseases. Meanwhile, the collection of symptom information mentioned in this method is conducive to updating the etiological information and summarizing the epidemic characteristics of influenza, providing further support for the early warning and prevention of influenza.

**Significance Statement:** We propose a new method for monitoring influenza epidemics by regularly reporting the health status of individuals and propose criteria for different levels of influenza alert severity. Our results validate the feasibility of this method, which can detect influenza timely and accurately and make early warning, and use health resources more effectively. The results of this study provide a new perspective for the surveillance of emerging infectious diseases and provide theoretical support for the cohort study of infectious diseases.

## 1. Introduction

Influenza, is an acute respiratory infection caused by the influenza virus^1^. Of all the pathogens of public health concern, influenza viruses are unique because of their ability to evolve and spread rapidly^2,3^. Generally, influenza causes mild illness, but it can also lead to severe illness and serious complications in some individuals^4^, with mortality rates typically highest in older populations and those with chronic and underlying medical conditions^5,6^.

Influenza causes a significant burden of morbidity and mortality^7^, estimated to be as many as half a million deaths per year worldwide^8^. It imposes a severe economic burden, which primarily includes unavoidable medical expenditures (direct costs) and lost productivity due to absenteeism from work (indirect costs)^4^. It is estimated that in the United States, influenza costs account for 65% of the total economic burden of all vaccine-preventable diseases^9^.Influenza is a major burden on human health and a major public health challenge^10^, which highlights the importance of rational and effective surveillance.

At the same time, many newly emerging infectious diseases, especially respiratory infectious diseases, are easy to break out in a wide range due to their rapid spread and strong infectivity^11^. These diseases may cause serious consequences, such as threatening life safety, undermining economic stability and even causing social unrest^12^. However, these respiratory infections are easily misdiagnosed as influenza in the early stages of diagnosis. This is due to the similarities in the signs and symptoms of respiratory virus infection and influenza virus infection^13^. For example, the common symptoms of COVID-19 are fever, cough, and runny nose^14^, which led to the initial misdiagnosis of COVID-19 as common influenza. Therefore, daily monitoring influenza and obtaining relevant data are of great significance for timely detection of new infectious diseases.

Because the onset, duration, intensity, and extent of transmission of influenza epidemics are unpredictable^15^, but the occurrence of new infectious diseases is inevitable, it is necessary to do routine surveillance as well as seasonal surveillance. Timely and accurate reporting of influenza epidemics can further reduce the health and economic burden, but current influenza disease surveillance faces many challenges. Since the beginning of the 21st century, most countries have established influenza surveillance system based on sentinel surveillance, hospital admission and mortality surveillance, but this system has limitations in the timeliness of information feedback and insufficient reliability of information^16^. Influenza sentinel hospitals do not have full coverage, some clinics or small hospitals do not report influenza cases, and even some influenza-infected persons do not seek medical attention, and some influenza cases cannot be confirmed by laboratory tests^17^,these are the shortcomings of the existing official monitoring system.

With the progress of surveillance technology and diagnosis and treatment, the surveillance system of influenza has been further developed. For example,a medical school in Japan has developed a influenza-tracking mobile app that uses volunteers to fill in a self-administered questionnaire to detect influenza activity early^18^. Compared with the official sentinel surveillance, it is more convenient and rapid, but the selection of participants is biased and cannot reflect the trend of influenza epidemic. There are also studies that use social media content related to influenza epidemic as indicators to monitor influenza epidemic, but this method has limitations in coverage and representativeness^19^. It can be seen that a perfect influenza surveillance method has not to be proposed. High-quality surveillance systems need to alert the public to preventive measures and health-care providers to advance medical preparedness, alert the public to preventive measures and health-care providers to advance medical preparedness, inform relevant decisions by policymakers, and prioritize resources for those most at risk of serious consequences, thereby reducing morbidity, mortality, and economic costs^5,20^. At the same time, surveillance systems also need to be able to detect influenza outbreaks and epidemics in a timely manner, and unusual increases in influenza cases can provide early warning of emerging infectious diseases.

In order to achieve timely warning and reasonable prevention, this paper proposes a regional influenza surveillance method based on fixed individuals which is based on reasonable statistical methods and threshold theory. The existing influenza-like illness (ILI) surveillance system is based on real-time influenza cases reported by sentinel hospitals, that is, only sick individuals are recorded in real time. However, the method proposed by us is to rationally select representative fixed monitoring individuals according to the epidemic characteristics of influenza, and make them directly upload their physical conditions on a regular basis to judge the occurrence of influenza. In our proposed surveillance system based on fixed individuals, relevant information can be directly uploaded in time after the monitoring object has symptoms, avoiding the deficiency that sentinel surveillance may not detect cases in time. The monitoring system sets the coverage range at the early stage of sampling and can represent the population with different characteristics, which avoids selection bias and makes the monitoring results more reliable. This monitoring system also involves different degrees of early warning of the severity of influenza, this provides a more accurate strategic direction for subsequent influenza prevention and control, and can make more reasonable use of medical and health resources.

Importantly, the surveillance method based on fixed individuals can provide long-term follow-up of the monitored subjects, which provides a theoretical basis for the cross-sectional study of infectious diseases and makes up the gap in the cohort study of infectious diseases. In addition, because other emerging respiratory infections have similar symptoms to influenza, there may be an unusual increase in influenza data before their outbreak. In December 2018, influenza activity in the European region continued to increase, with influenza trends up to about 50%. And before COVID-19 was identified, the incidence of influenza increased from the previous year can be also found in places such as China and Brazil^21,22^. Therefore, the monitoring of influenza will play an important role in the monitoring and early warning of new infectious diseases. The regional influenza surveillance method based on fixed individuals proposed in this paper can detect influenza more accurately and timely and give early warning, and make more effective use of health resources, which is of great significance for the development of influenza surveillance system.

## 2. Method

### 2.1 Data sources

A new monitoring method is proposed in this study, but this method has not been put into practice. To prove the feasibility of this method, we present a case. The data in the case study were generated based on surveillance data from the Centre for Disease Control and Prevention (CDC) in Jilin Province, China. If the system is put into use, it can be used as a data source based on the data reported by fixed individuals collected by the system.

### 2.2 Monitoring method

The first step is to summarize the characteristics of the influenza epidemic.

(1) Age difference: the age group under 15 years old is the main susceptible group^23,24^;
(2) Sex differences: sex differences in influenza prevalence vary with age. In the adolescent group, males are more likely to be infected with the flu virus. In the young and middle-aged group, women are generally more vulnerable to the severe consequences of viral infection. In the older age group, men were more likely to develop influenza illness than women^25-27^;
(3) Occupational differences: during the influenza epidemic, people who go out more or (and) are in contact with infected people have more susceptibility^28^ (health care workers^29^, students, etc.);
(4) Seasonal: The Northern Hemisphere and the southern hemisphere have different characteristics of influenza epidemic due to their different geographical locations and climatic conditions^30,31^. Influenza cycles vary depending on geographic location, virus type, and climatic conditions^30^.

In the second step, the sample size is estimated, and the stratified sample size is estimated and the fixed individual is extracted according to the distribution characteristics and influencing factors. Number the fixed individuals and have them complete the basic information form, as shown in Table S1.

The sampling method is stratified sampling, and the specific implementation method is as follows.

S1. The overall sample size (*N*) is estimated at the province / city, and the calculation formula is as follows.

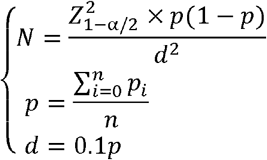

where *Z*_1−*α/2*_ is a two-sided test, which is 1.96 when *α* taking 0.05. *p* is the case rate. *p*_*i*_ is average prevalence of the non-epidemic period i. *n* is total non-epidemic period, *d* is the admissible error.

S2. Combined with the epidemic characteristics, with age, gender and occupation as stratification factors, the overall population is divided into non-overlapping layers, and the number of individuals to be selected in each layer is determined according to the proportion. The proportion is determined as follows.

a. 0∼14 years old accounted for 60% of the total, 15∼59 years old accounted for 35%, over 60 years old age accounted for 5%;
b. In all age, men accounted for 55% and women accounted for 45%;
c. In the age group of 0 to 14, the sampling proportion of preschool children is lower than that of school-age children and adolescents; in 15∼59 people according to occupational stratification, medical personnel; participants in large activities; the sampling proportion of vulnerable people and employees in pension institutions, long-term care institutions and welfare institutions is higher than that of the general population;

S3. Draw a corresponding number of fixed individuals in each layer by simple random sampling;

S4. The fixed individuals drawn from each layer are merged to obtain the overall sample.

#### The third step is to determine the monitoring plan

According to the regional climate characteristics, the preliminary surveillance upload plan of different regions is determined, and then the preliminary upload plan is adjusted according to the existence and characteristics of seasonal influenza epidemics in the region, and the final annual upload plan is determined.

Based on the literature study and the special climate characteristics of the detail areas, the epidemiological seasons in each region were determined, and the uploading cycle was more compact during the epidemic season and could be appropriately relaxed during the non-epidemic season. During the epidemic period, the upload plan is every Monday and every Thursday, that is, upload twice a week. This kind of reporting schedule mainly considers that the average incubation period of influenza is 3-7 days, and the frequency of twice a week can fully guarantee the real-time data. In areas with more than two epidemiological seasons in a year, the non-epidemic period is still uploaded every Monday, while areas with two or fewer epidemiological seasons in a year can upload fortnightly, i.e. every other Monday.

#### The fourth step is the uploading of monitoring results

Fixed individuals will upload influenza pathogen test results and symptoms according to the annual upload plan. First, the monitored subjects are trained in the correct methods of temperature detection, pathogen self-testing, and uploading of results. At the same time, the monitored objects are notified of the specific time of the annual upload plan to ensure that the individual upload results on time. In addition, it is necessary to issue test reagents to monitored personnel regularly to ensure the smooth monitoring. If the monitored subjects do not show flu-like symptoms, the monitoring results should be uploaded regularly according to the upload plan. However, if the monitoring object has symptoms suspected of influenza, the relevant symptom information should be reported immediately (see Table S2), and the information should be reported continuously before the symptoms disappear, and the daily uploading and filling can be resumed after the symptoms disappear.

The collection of symptom information can first provide support information for pathogen typing analysis, which can better assist the early warning of influenza epidemic and the update of etiological information. Second, symptom information can be used to summarize the characteristics of this influenza epidemic and provide better guidelines for future influenza prevention and control. Finally, the collection of information can provide more detailed data for future infectious disease cohort studies.

It should be noted that the upload schedule interval is different, so the data in the table needs to be adjusted by the correction coefficient. The specific adjustment method is as follows.

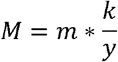

where *M* is the number of corrected cases. *m* is the number of uploaded cases. *y* is the days apart from the last upload. *k* is the maximum upload interval number of days.

#### The fifth step is to determine the threshold theory and analyze data

The incidence of influenza at the n-th upload was compared with the average incidence of *n* − 1, *n*− 2 and *n* − 3 upload, and χ^2^ test was used to compare the incidence.

Based on the real data over the past years, the early warning value of influenza epidemic is determined and the corresponding threshold theory is given. The influenza incidence rate for the current upload (*n* times), when compared to the average incidence rate for the previous three times (*n* − 1, *n*− 2, *n* − 3 times), if it is a decline indicates that there is no risk of pandemic for the time being, if it is an increase, then the uploaded influenza incidence and the average of the previous three times of the average incidence of influenza statistically test, the test is a two-sided test, *α* take 0.05, using χ^2^ distribution table can be seen the bounding value is 3.96, when the value of the test χ^2^ is greater than 3.96, it means that the difference time uploaded influenza incidence compared to the average number of incidence of *n* + 3, is statistically significant, indicating the possibility of an influenza epidemic. When the *n* times influenza incidence suggests that there is a possibility of influenza epidemic, the *n* − 1, *n* − 2, *n* − 3times, the judgment standard is the same as that of the *n* times, when the two consecutive comparisons are up and there are significant differences, suggesting that there is a high probability of influenza outbreaks, and the need to report the data, and will be consecutively there is a difference between the two times of the data, respectively define the two consecutive differences as the *H*_*1*_ and *H*_*2*_ of this epidemic.

The uploaded data is revised and tested. When the test results indicate that there is a high possibility of influenza epidemic, the results will be reported, and the reported data will be compared with the data of the same period over the past years.

Previous data of the same period are defined as follows. During the statistical testing of the uploaded data during the previous annual monitoring, When there are differences between the two continuous tests, These two times are defined as 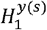 and 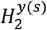, respectively (where year is the year, season refers to the prevalent season, if there is a merger in the popular season, write the year in the form of a time period. The epidemic season is based to the beginning of the epidemic). Matching the historical data to the current annual data by the epidemic season, Data for 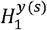 and 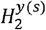 of the previous epidemic season are defined as data of the same period for *H*_*1*_ and *H*_*2*_ of the monitoring year. All previous 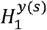 and 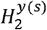 data are added and averaged respectively, namely, the average of the previous epidemic period (if some historical period data does not exist, then average the years after elimination). After the data report, compare the data of *H*_*1*_ and *H*_*2*_ times respectively with the average data of the previous epidemic period (through the seasonal matched 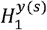 and 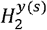). If the number of cases is all significantly higher than the previous same period, the first level warning is triggered; only one higher than the previous period triggers secondary warning; and no statistical difference from the previous data, or below the same period in the previous years triggering the third level of warning.

Effective prevention and control measures are needed to report the data simultaneously and start the corresponding level of early warning. Combined with the threshold theory, the early warning and lifting of the early warning should be carried out in time, and the prevention and control measures should be formulated for the influenza epidemic during the early warning period.

The threshold theory for lifting the early warning. Trigger early warning after correction data compared with epidemic period average onset data, when the χ^2^ value is less than 3.96, when there is no statistical difference warning, or χ^2^ value greater than 3.96, there are statistical differences between the two, but the upload data below the epidemic period average onset data, can remove warning, restore to daily monitoring, if the upload data is higher than the epidemic period average onset data, and the comparison exist statistical differences, continuous prevention and control, strengthen the monitoring. For the non-epidemic period average incidence rate is calculated as follows: the actual epidemic period (i.e., the period from the beginning of the warning to the lifting of the warning) in the previous annual monitoring cycle is excluded, and the remaining is defined as a non-epidemic period. Because there are differences in the number of days between reports, the data uploaded in the non-epidemic period are corrected using correction coefficients and then averaged.

## 3. Example

We choose Jilin Province of China as the basis of illustration. According to the Qinling-Huaihe River line in China, this area belongs to northern China and has the characteristics of “influenza epidemic period occurs mostly in the cold season of winter and spring”. We carried out the implementation of the example according to the above conditions, and in order to make the example more realistic, the data were generated based on the experimental data of influenza in Jilin Province from Jun.30, 2017 to Jun.30, 2018. The data for the same period in previous years were generated based on influenza data from 2012 to 2016 in Jilin Province.

The first step, the Chinese region of Jilin Province epidemic characteristics and the characteristics of the above mentioned is similar. Among them, the region is located in northern China, there are two epidemics of influenza in winter and spring, and most of them are prevalent in winter.

In the second step, the sample size is estimated, and the stratified sample size is estimated and the fixed individual is extracted according to the distribution characteristics and influencing factors. Number the fixed individual and have it fill out the basic situation table.

S1. The prevalence estimates are proxied using the influenza incidence rate, which can be somewhat representative of prevalence because influenza is a short-term disease. The final combined influenza 2017 and 2018 incidence rate will be set at 5 per 10,000 population^32^.The final calculated overall sample size (*N*) estimate is shown below.

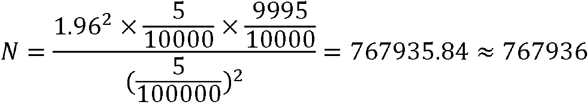

S2. Combined with the epidemic characteristics, the overall population is divided into non-intersecting strata using age, sex, and occupation as stratification factors, and the number of individuals to be sampled from each stratum is determined proportionally, with the proportions determined as follows.

a. 60% of the total number of individuals in the age group 0-14 years, 35% in the age group 15-59 years, and 5% in the age group above 60 years;
b. 55% males and 45% females in each age stratum;
c. In the 0 to 14 age group, the sampling proportion of children in early childhood is lower than that of school-age children and adolescents; in the secondary stratification according to occupation in the 15 to 59 age group, the sampling proportions of medical personnel; participants in large-scale activities and security personnel; and vulnerable people and employees of places where people gather, such as elderly care institutions, long-term care institutions, and welfare homes, are higher than those of the general population;

S3. Simple random sampling is used to draw a corresponding number of fixed individuals in each stratum;

S4. Combine the fixed individuals drawn from each stratum to obtain the overall sample. The final sampling results are shown in Figure 1.

**Figure 1.**
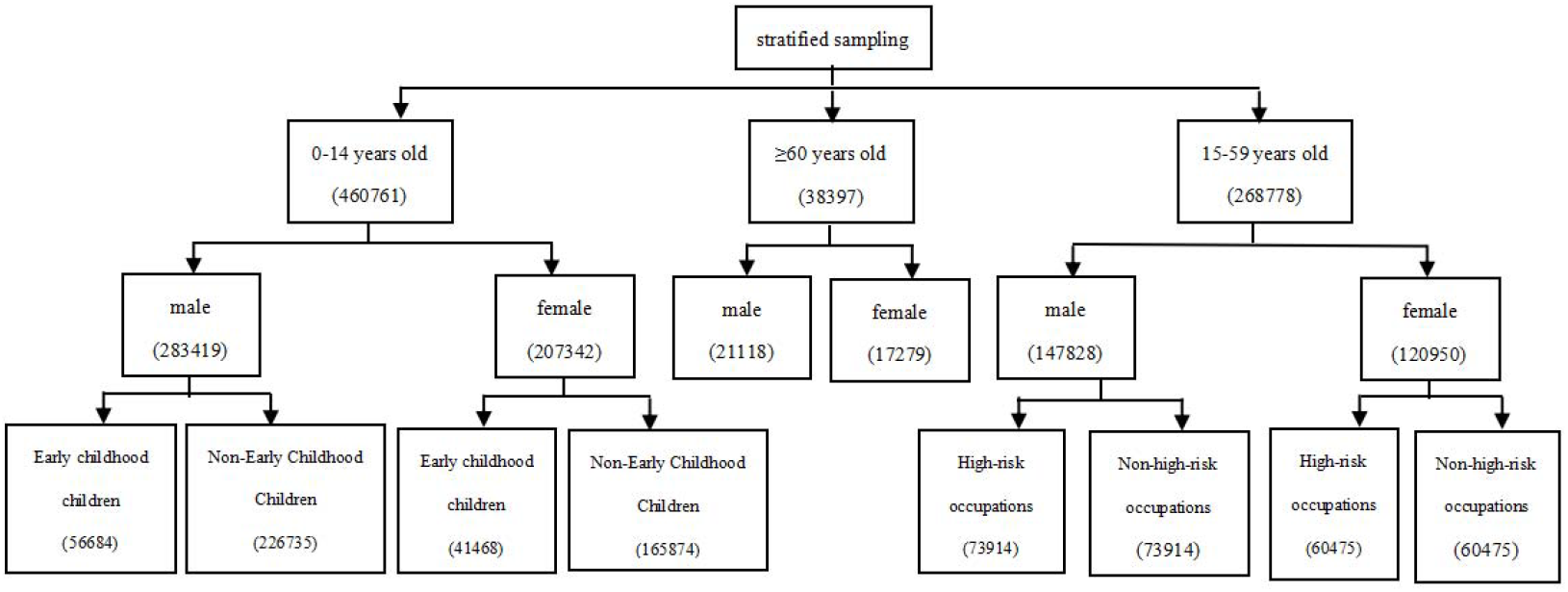
Final sampling result.

The third step is to determine the upload monitoring plan of Jilin Province.

The initial upload plan is determined according to the climate characteristics of Jilin Province. Jilin Province is a northern province, and existing studies are consistent with the finding that “influenza epidemics tend to occur during the cold winter and spring seasons”. Therefore, the uploading plan is focused on the winter and spring seasons.

Then, studies showed that there was a seasonal influenza epidemic in the region. Because of the early winter in Jilin Province, early warning, prevention and control will obtain better benefits. According to relevant research and historical data of Jilin Province, the epidemic period is determined to be 50th week to 6th week of the following year in, and 10th week to 13th week in spring. Adjust the initial upload plan based on the above content and determine the final annual upload plan. CDC surveillance data uploaded every Monday and Thursday are shown in the Table S3.

The results show that the number of people uploaded during the non-epidemic period is relatively stable, and Jilin Province is a very representative northern province, so it is sufficient to upload the data once every two weeks during the non-epidemic period, and the uploading plan during the epidemic period is set to upload the data on Mondays and Thursdays, uploading twice a week for each individual, and the specific uploading dates are shown in Table S4.

The fourth step is for fixed individuals to upload influenza pathogen test results and symptoms according to the annual upload schedule.

Individuals are trained in advance, including normal body temperature detection, pathogen self-testing, and results uploading methods. Inform the monitoring object of the specific time of the annual upload plan to ensure that it can upload the results normally. In addition, additional pathogen self-testing reagents need to be issued regularly. The monitoring subjects regularly upload the monitoring results according to the monitoring plan, but the monitoring subjects need to fill in the information form after symptoms appear until the symptoms disappear. Take monitoring individual No.52 as an example, the results were regularly uploaded according to the monitoring plan. Surveillance individual No.52 developed influenza-like symptoms on Jan.03,2018, and her symptoms disappeared on Jan.10,2018 Therefore, the monitoring individual No.52 needs to continuously fill in and report information from Jan.03, 2018 to Jan.10, 2018, and continue to upload monitoring results according to the monitoring plan after Jan.10,201

It should be noted that the upload progress interval is different, and the number of uploaded cases (*m*) needs to be adjusted by the correction coefficient to get the corrected number of cases (*M*), and then the data can be analyzed. Jilin is a northern province. Therefore, the value of *K* (maximum number of upload days) is 14, and the value of y (the days apart from the last upload) varies with the reporting date. The value of y is 3,4, or 14. The specific adjustment methods are as follows.

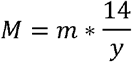

The fifth step is data analysis

First, the warning value of influenza epidemic is determined, then, the corresponding threshold theory is given based on the real data of the past years, and the data correction and testing of the uploaded data is carried out. The statistics of the number of incidences on each upload date are shown in Table S5. Based on the threshold theory and data analysis results, timely and reasonable early warning should be issued, the early warning should be lifted, and timely prevention and control measures should be formulated for the influenza epidemic after the early warning.

The results show that the number of uploaded cases in Dec.11,2017 is 347, and the number of corrected cases is 694. The corrected cases show an increasing trend, and their χ^2^ value is calculated as 31.195 > 3.96, which means that the uploaded cases of influenza are distributed differently compared to the previous average cases, and according to the threshold theory, there is a possibility of an influenza pandemic.

The number of uploaded cases on Dec.14,2017 is 157, and the number of corrected cases is 733. The corrected incidence shows an upward trend, and its χ^2^ value is calculated as 43.653 > 3.96, which means that the uploaded influenza incidence has distributional differences compared with the previous average incidence, and according to the threshold theory, this upload triggers an early warning and requires effective preventive and control measures.

Report the results when it is suggested that there is a high probability of an influenza epidemic, and at the same time compare the reported data with the same period of the previous years. According to the results, activate the corresponding warning level.

Compare the reported data with data from the same period in previous years, and a combined epidemic season has occurred in this upload year. For Dec.11,2017 (the first), the average number of uploaded cases in its previous simultaneous epidemic period is 150, and the average number of corrected cases is 700. Its value is calculated as 0.026 < 3.96, which means that there is no difference in the distribution of this uploaded influenza incidence compared to the average incidence of the historical contemporaneous epidemics, and for Dec.14,2017 (the first time), its average number of uploaded cases in the previous contemporaneous period is 169, and the average number of corrected cases is 789. Calculate χ^2^ value as 2.062 < 3.96, which means that there is no distributional difference between the uploaded influenza cases and the average cases of previous simultaneous epidemics; thus, it can be seen that there is no significant increase in the two times compared to the previous years, and therefore, the third level of warning is activated.

After the early warning, comparing the latest uploaded number of cases with the average number of cases in the non-epidemic period, the χ^2^ value is consistently greater than 3.96, and the two epidemics in Jilin Province are close to each other, so the warning is never lifted. Until Apr.16,2018 when the number of uploaded cases is 438 and the number of corrected cases is 438.The revised incidence situation showed a decreasing trend, and its value is calculated as 3.55 < 3.96, i.e., it means that the uploaded influenza incidence rate is not statistically different from the previous average incidence rate. According to the threshold theory, i.e., the alert is lifted on Apr.16,2018 for daily surveillance.

From the beginning of the alert, the following preventive and control measures are taken in the influenza epidemic areas.

(1) Health education. dissemination of information on influenza prevention and control and behavioral interventions for residents of Province A. Health promotion agencies at all levels help individuals and groups acquire knowledge of health care and establish a concept of health;
(2) Health promotion. stipulating the respective responsibilities of individuals and society for health, and uniting relevant departments as well as the government to pay attention to individual health;
(3) Vaccination. arranging influenza vaccination for people of high influenza-prone ages and occupations, and arranging special vaccination lanes for elderly people with basic illnesses and pregnant women with low resistance to prevent influenza virus infection;
(4) Rational allocation of medical resources. Residents of Province A should be reminded to prepare medicines, including some common cough medicines and fever-reducing medicines, etc. major hospitals should increase fever outpatient clinics; and, at the same time, increase beds in departments related to influenza complications, such as the Infectious Diseases Department (lung infections) and Cardiology Department (influenza viral myocarditis); and at the same time, establish an expert group on medical prevention, control, and treatment;
(5) Timely, open, and transparent information and good communication. Provide the community with timely and up-to-date information about the influenza pandemic to minimize public panic;
(6) Increase social distance. If necessary, isolate and close schools, cancel gatherings, and restrict travel in influenza epidemic areas in Jilin Province;
(7) Enhance surveillance. Change the upload schedule from once every two weeks to twice a week during the non-epidemic peak period to detect, diagnose, report, and treat cases in a timely manner.

## 4. Discussion

Influenza is an acute respiratory illness caused by the influenza virus. Influenza virus has the characteristics of strong variability, fast transmission speed, and strong infective. It can occur on a large scale in specific seasons and in crowded places^33^. Globally, influenza epidemics have a significant impact on the public health burden^34^. There have been five influenza pandemics since the beginning of the 20th century. the 1918 Spanish flu, the 1957 Asian flu, the 1968 Hong Kong flu^35^, the 1977 Russian flu^36^, and the 2009 H1N1 flu^37^. These pandemics have resulted in hundreds of millions of infections and millions of deaths^38^. Due to the burden of influenza disease and the threat of influenza pandemic, a complete influenza surveillance system is required.

Sentinel surveillance is now the main method of monitoring influenza prevalence in most countries^39^. The literature shows that sentinel surveillance has a lag, and the detection of influenza is not timely and sensitive enough^40^. In this study, a regional influenza surveillance method based on fixed individuals is proposed. In this method, individuals are used as the monitoring points, and the monitoring subjects are required to fill in the relevant information regularly according to the unified monitoring method, making it more convenient to detect the presence of influenza. At the same time, sentinel monitoring needs to be fixed point, timing, quantitative detection, there will be a shortage of resources. Our proposed method determines the surveillance plan according to the characteristics of regional and seasonal changes of influenza. Compared with the previous surveillance methods, it is more economical in manpower and material resources and more rational in the use of health resources.

Some researchers have proposed to monitor and predict influenza epidemics based on search engine data. This method may misjudge influenza by relying on search data and may be difficult to implement in different countries^41^. Our individual-based approach to influenza surveillance is representative and allows for better prediction of influenza epidemics. Zhen Yang et al. used face recognition collection and thermal imaging measurement technology to monitor the absenteeism rate and body temperature of studying students and implemented the influenza school infectious syndrome monitoring system^42^. The system is simple to operate and low cost, but its accuracy needs to be improved. Moreover, the monitoring system is only implemented in the student population, without monitoring the whole population, and lacks the representation of the whole population. In Japan, influenza outbreaks and epidemics are monitored by estimating the number of influenza cases through the number of prescriptions for anti-influenza drugs^40^. During the flu season, anti-flu drugs are often used to prevent the flu. In addition, in the case of influenza outbreaks, media coverage and public panic may cause people to buy and stock up on anti-influenza drugs. These may lead to an overestimation of the total number of influenza cases, resulting in a misestimation of the occurrence of influenza. Our approach avoids these drawbacks from the beginning of design sampling. According to the characteristics of influenza epidemic, we conducted stratified sampling and selected people with different characteristics as representatives to achieve full coverage of the population and increase the reliability of surveillance results. At the same time, we monitor and report the symptoms of influenza, and use pathogen detection to check whether there is disease. Thus, the accuracy of monitoring will be greatly improved, and the occurrence of influenza can be predicted more accurately.

The symptoms and signs of many respiratory infectious diseases are similar to those of influenza. Diseases transmitted by the upper respiratory tract can easily cause widespread infection in a short period of time, the surveillance system proposed by us is based on symptoms to monitor diseases, which can have a certain suggestive effect on the occurrence of respiratory infectious diseases other than influenza. Thus, the influenza surveillance system plays a sentinel role in monitoring emerging infectious diseases, enabling timely detection of infectious diseases and regular epidemic prevention and control. At the same time, our surveillance is ongoing and surveillance data is always uploaded regularly, so we are able to obtain the full trajectory of the disease epidemic and follow-up cohort study data, which provides the basis for cohort studies of infectious diseases. The monitoring of infectious diseases in a certain period of time also provides more support for cross-sectional studies of infectious diseases.

This study identifies fixed and regularly monitored individuals through full coverage sampling, guides these individuals, and can accurately and effectively collect personal health data. Then, pathogen detection data of symptomatic individuals could be used to determine whether there is a disease epidemic. If a symptomatic individual fails to detect a specific infection from a known pathogen, it may indicate the emergence of a new disease. This operation is operated at a lower cost and improves the sensitivity and accuracy of the monitoring system. In addition, we put forward an innovative basis for the classification of influenza prevalence, considering the influenza data of the same period and the historical data of previous years to assess the severity of influenza epidemic. This provides a reference for the prevention and control of influenza. This monitoring method can simultaneously collect relevant epidemiological information, used to reflect the epidemiological changes of influenza, assess the intensity of influenza, and provide a basis for the development of prevention strategies.

In order to verify the feasibility of this surveillance method, we used the influenza data of a region to monitor the occurrence of influenza in the region, and the results showed that the method could accurately detect the existence of influenza and judge its epidemic grade. At the same time, the method can make reasonable and effective use of health resources, collect relevant epidemiological information, and provide a basis for preventing and controlling the epidemic of influenza. This method has not been widely used, but it provides a framework to monitor and warn the epidemic of influenza.

## 5. Conclusions

This study provides a regional influenza surveillance method based on stationary individuals. This method can effectively monitor the occurrence of influenza and act as a sentinel for the discovery of some unknown new infectious diseases. At present, there is no cohort study database for infectious diseases, and this fixed individual surveillance method, once put into use, can not only provide material for cross-sectional studies but also fill the gap in infectious disease cohort studies. At the same time, the collection of symptom information can further provide data support for the update of etiology information, the summary of influenza epidemic characteristics and the cohort study. We also validated the feasibility of the method using regional data, effectively proving that the surveillance method proposed in this study can accurately detect the presence of influenza. Importantly, the framework proposed in this study is universal and can be arbitrarily extended to the surveillance and early warning work of other infectious diseases.

## Data Availability

All data produced in the present study are available upon reasonable request to the authors
All data produced in the present work are contained in the manuscript

## Supplementary Material

Supplementary material is available at PNAS Nexus online.

## Acknowledgments

Not applicable

