## Supplementary material for "New framework for the surveillance and early warning of influenza: fixed individuals regular reporting mechanism": Table S1-S5

Table S1 Monitoring individual basic information table

| **Monitoring individual basic information table** | | | |
| --- | --- | --- | --- |
| ID: ☐ | | | |
| 1. Basic situation | | | |
| Name |  | Gender | ☐Male ☐Female |
| Age |  | Contact number |  |
| Height (cm) |  | Weight (kg) |  |
| Current address: | | | |
| Occupation：  ☐ Agriculture, Forestry, Fishing ☐ Health Care, Social Assistance  ☐ Management, Administrative ☐ Teacher  ☐ Student ☐Lawyer  ☐ Other items: | | | |
| 2. Past medical history | | | |
| 2.1 Are there any of the following underlying diseases? | | | |
| (1) Chronic lung diseases   ☐Yes ☐No ☐Not clear  If yes, ☐asthma ☐chronic bronchitis ☐emphysema ☐chronic obstructive pulmonary disease ☐Obstructive sleep apnea syndrome ☐Other (please fill in disease name): | | | |
| (2) Cardiovascular diseases ☐Yes ☐No ☐Not clear  If yes, ☐hypertension ☐coronary heart disease ☐other (please fill in the name of the disease): | | | |
| (3) Metabolic diseases ☐Yes ☐No ☐Not clear  If yes, ☐diabetes (please select the type of diabetes: ☐type 1 ☐type 2 ☐not clear) ☐hyperlipidemia ☐Other (Please fill in disease name): | | | |
| (4) Chronic kidney disease ☐Yes ☐No ☐Not clear  If yes, please fill in the name of the disease: | | | |
| (5) Chronic liver diseases  ☐Yes ☐No ☐Not clear  If yes, please fill in the name of the disease: | | | |
| (6) Cancer/tumor ☐Yes ☐No ☐Not clear  If yes, please fill in the name of the disease: | | | |
| (7) Are there any other systemic diseases  ☐Yes ☐No ☐Not clear  ☐ Other diseases 1:  ☐ Other diseases 2:  ☐ Other diseases 3: | | | |
| 2.2 Have you been vaccinated against seasonal flu in the past year?  ☐Yes ☐No ☐Not clear | | | |

Table S2 Report information table

| **Report information table** | | |
| --- | --- | --- |
| 1. Basic information | | |
| ID |  | Reporting date |
| 2. Clinical manifestation | | |
| 2.1 Have any flu-like symptoms?  ☐Yes ☐No ☐Not clear | | |
| 2.2 Symptom filling | | |
| (1) Fever ☐Yes ☐No ☐Not clear  Please specify the maximum body temperature after the onset of this disease ℃ | | |
| (2) Sore throat ☐Yes ☐No ☐Not clear | | |
| (3) Cough ☐Yes ☐No ☐Not clear | | |
| (4) Shortness of breath or difficulty breathing ☐Yes ☐No ☐Not clear | | |
| (5) Shortness of breath ☐Yes ☐No ☐Not clear  Please specify that the highest respiratory rate observed after this episode:  times/min | | |
| 3. Contact history | | |
| (1) Have you been vaccinated against seasonal flu in the past year?  ☐Yes ☐No ☐Not clear | | |
| (2) Have you had contact with dead livestock?  ☐Yes ☐No ☐Not clear | | |
| (3) Have you been in contact with similar cases?  ☐Yes ☐No ☐Not clear | | |
| 4. Pathogen self-test results | | |
| ☐A(H1N1) ☐A(H3N2) ☐A(H1N1) ☐ No subtypes ☐ B  ☐ Mixed type (type/subtype) ☐ Not clear | | |

Table S3 Upload CDC surveillance data on Mondays and Thursdays

| Reporting date | Epidemic peak | Number of reported | Reporting date | Epidemic peak | Number of reported |
| --- | --- | --- | --- | --- | --- |
| 2017.07.03 | No | 93 | 2018.01.01 | Yes | 197 |
| 2017.07.06 | No | 75 | 2018.01.04 | Yes | 150 |
| 2017.07.10 | No | 99 | 2018.01.08 | Yes | 200 |
| 2017.07.13 | No | 76 | 2018.01.11 | Yes | 169 |
| 2017.07.17 | No | 101 | 2018.01.15 | Yes | 225 |
| 2017.07.20 | No | 85 | 2018.01.18 | Yes | 177 |
| 2017.07.24 | No | 112 | 2018.01.22 | Yes | 236 |
| 2017.07.27 | No | 82 | 2018.01.25 | Yes | 169 |
| 2017.07.31 | No | 109 | 2018.01.29 | Yes | 225 |
| 2017.08.03 | No | 76 | 2018.02.01 | Yes | 162 |
| 2017.08.07 | No | 101 | 2018.02.05 | Yes | 216 |
| 2017.08.10 | No | 86 | 2018.02.08 | Yes | 142 |
| 2017.08.14 | No | 115 | 2018.02.12 | No | 169 |
| 2017.08.17 | No | 83 | 2018.02.15 | No | 119 |
| 2017.08.21 | No | 111 | 2018.02.19 | No | 158 |
| 2017.08.24 | No | 91 | 2018.02.22 | No | 121 |
| 2017.08.28 | No | 118 | 2018.02.26 | No | 155 |
| 2017.08.31 | No | 81 | 2018.03.01 | No | 127 |
| 2017.09.04 | No | 108 | 2018.03.05 | Yes | 169 |
| 2017.09.07 | No | 86 | 2018.03.08 | Yes | 121 |
| 2017.09.11 | No | 111 | 2018.03.12 | Yes | 161 |
| 2017.09.14 | No | 84 | 2018.03.15 | Yes | 120 |
| 2017.09.18 | No | 106 | 2018.03.19 | Yes | 160 |
| 2017.09.21 | No | 83 | 2018.03.22 | Yes | 122 |
| 2017.09.25 | No | 110 | 2018.03.26 | Yes | 163 |
| 2017.09.28 | No | 89 | 2018.03.29 | Yes | 111 |
| 2017.10.02 | No | 111 | 2018.04.02 | No | 148 |
| 2017.10.05 | No | 91 | 2018.04.05 | No | 97 |
| 2017.10.09 | No | 107 | 2018.04.09 | No | 129 |
| 2017.10.12 | No | 87 | 2018.04.12 | No | 91 |
| 2017.10.16 | No | 109 | 2018.04.16 | No | 121 |
| 2017.10.19 | No | 89 | 2018.04.19 | No | 90 |
| 2017.10.23 | No | 105 | 2018.04.23 | No | 114 |
| 2017.10.26 | No | 88 | 2018.04.26 | No | 87 |
| 2017.10.30 | No | 113 | 2018.04.30 | No | 115 |
| 2017.11.02 | No | 82 | 2018.05.03 | No | 88 |
| 2017.11.06 | No | 102 | 2018.05.07 | No | 117 |
| 2017.11.09 | No | 92 | 2018.05.10 | No | 89 |
| 2017.11.13 | No | 110 | 2018.05.14 | No | 119 |
| 2017.11.16 | No | 90 | 2018.05.17 | No | 86 |
| 2017.11.20 | No | 115 | 2018.05.21 | No | 115 |
| 2017.11.23 | No | 94 | 2018.05.24 | No | 91 |
| 2017.11.27 | No | 109 | 2018.05.28 | No | 112 |
| 2017.11.30 | No | 95 | 2018.05.31 | No | 90 |
| 2017.12.04 | No | 104 | 2018.06.04 | No | 120 |
| 2017.12.07 | No | 149 | 2018.06.07 | No | 87 |
| 2017.12.11 | Yes | 198 | 2018.06.11 | No | 118 |
| 2017.12.14 | Yes | 157 | 2018.06.14 | No | 78 |
| 2017.12.18 | Yes | 210 | 2018.06.18 | No | 105 |
| 2017.12.21 | Yes | 160 | 2018.06.21 | No | 84 |
| 2017.12.25 | Yes | 214 | 2018.06.25 | No | 112 |
| 2017.12.28 | Yes | 148 | 2018.06.28 | No | 87 |

Table S4 Specific upload schedule

| Reporting date | Epidemic peak | Remark | Reporting date | Epidemic peak | Remark |
| --- | --- | --- | --- | --- | --- |
| 2017.07.03 | No | Monday | 2018.01.22 | Yes | Monday |
| 2017.07.17 | No | Monday | 2018.01.25 | Yes | Thursday |
| 2017.07.31 | No | Monday | 2018.01.29 | Yes | Monday |
| 2017.08.14 | No | Monday | 2018.02.01 | Yes | Thursday |
| 2017.08.28 | No | Monday | 2018.02.05 | Yes | Monday |
| 2017.09.11 | No | Monday | 2018.02.08 | Yes | Thursday |
| 2017.09.25 | No | Monday | 2018.02.12 | No | Monday |
| 2017.10.09 | No | Monday | 2018.02.26 | No | Monday |
| 2017.10.23 | No | Monday | 2018.03.05 | Yes | Monday |
| 2017.11.06 | No | Monday | 2018.03.08 | Yes | Thursday |
| 2017.11.20 | No | Monday | 2018.03.12 | Yes | Monday |
| 2017.12.04 | No | Monday | 2018.03.15 | Yes | Thursday |
| 2017.12.11 | Yes | Monday | 2018.03.19 | Yes | Monday |
| 2017.12.14 | Yes | Thursday | 2018.03.22 | Yes | Thursday |
| 2017.12.18 | Yes | Monday | 2018.03.26 | Yes | Monday |
| 2017.12.21 | Yes | Thursday | 2018.03.29 | Yes | Thursday |
| 2017.12.25 | Yes | Monday | 2018.04.02 | No | Monday |
| 2017.12.28 | Yes | Thursday | 2018.04.16 | No | Monday |
| 2018.01.01 | Yes | Monday | 2018.04.30 | No | Monday |
| 2018.01.04 | Yes | Thursday | 2018.05.14 | No | Monday |
| 2018.01.08 | Yes | Monday | 2018.05.28 | No | Monday |
| 2018.01.11 | Yes | Thursday | 2018.06.11 | No | Monday |
| 2018.01.15 | Yes | Monday | 2018.06.25 | No | Monday |
| 2018.01.18 | Yes | Thursday | — | — | — |

Table S5 The statistics of the number of incidences on each upload date

| Reporting date | Number of reported | Number of corrections | Average number of the first three times | $\chi^{2}$value |
| --- | --- | --- | --- | --- |
| 2017.07.03 | 93 | 326 | — | — |
| 2017.07.17 | 351 | 351 | — | — |
| 2017.07.31 | 388 | 388 | 355 | 1.466 |
| 2017.08.14 | 378 | 378 | 372 | 0.048 |
| 2017.08.28 | 403 | 403 | 390 | 0.213 |
| 2017.09.11 | 386 | 386 | 389 | 0.012 |
| 2017.09.25 | 383 | 383 | 391 | 0.083 |
| 2017.10.09 | 398 | 398 | 389 | 0.103 |
| 2017.10.23 | 390 | 390 | 390 | 0.000 |
| 2017.11.06 | 385 | 385 | 391 | 0.046 |
| 2017.11.20 | 407 | 407 | 394 | 0.211 |
| 2017.12.04 | 402 | 402 | 398 | 0.020 |
| 2017.12.11 | 347 | 694 | 501 | 31.195 |
| 2017.12.14 | 157 | 733 | 610 | 43.653 |
| 2017.12.18 | 210 | 735 | 721 | 110.179 |
| 2017.12.21 | 160 | 747 | 738 | 116.592 |
| 2017.12.25 | 214 | 749 | 744 | 117.673 |
| 2017.12.28 | 148 | 691 | 729 | 87.735 |
| 2018.01.01 | 197 | 690 | 710 | 87.245 |
| 2018.01.04 | 150 | 700 | 694 | 92.183 |
| 2018.01.08 | 200 | 700 | 697 | 92.183 |
| 2018.01.11 | 169 | 789 | 730 | 139.941 |
| 2018.01.15 | 225 | 788 | 759 | 139.369 |
| 2018.01.22 | 236 | 826 | 813 | 161.585 |
| 2018.01.25 | 169 | 789 | 814 | 139.941 |
| 2018.01.29 | 225 | 788 | 801 | 139.369 |
| 2018.02.01 | 162 | 756 | 778 | 121.480 |
| 2018.02.05 | 216 | 756 | 767 | 121.480 |
| 2018.02.08 | 142 | 663 | 725 | 74.397 |
| 2018.02.12 | 169 | 592 | 670 | 44.356 |
| 2018.02.26 | 553 | 553 | 603 | 30.500 |
| 2018.03.05 | 296 | 592 | 579 | 44.356 |
| 2018.03.08 | 121 | 565 | 570 | 34.543 |
| 2018.03.12 | 161 | 564 | 574 | 34.198 |
| 2018.03.15 | 120 | 560 | 563 | 32.834 |
| 2018.03.19 | 160 | 560 | 561 | 32.834 |
| 2018.03.22 | 122 | 570 | 563 | 36.287 |
| 2018.03.26 | 163 | 571 | 567 | 36.640 |
| 2018.03.29 | 111 | 518 | 553 | 19.919 |
| 2018.04.02 | 148 | 518 | 536 | 19.919 |
| 2018.04.16 | 438 | 438 | 491 | 3.549 |
| 2018.04.30 | 406 | 406 | 454 | 2.681 |
| 2018.05.14 | 413 | 413 | 419 | 0.043 |
| 2018.05.28 | 404 | 404 | 408 | 0.020 |
| 2018.06.11 | 415 | 415 | 411 | 0.019 |
| 2018.06.25 | 379 | 379 | 399 | 0.514 |
